# Perceived barriers to cervical cancer screening and motivators for at-home human papillomavirus self-sampling during the COVID-19 pandemic: Results from a telephone survey

**DOI:** 10.1101/2022.11.20.22282562

**Authors:** Susan L. Parker, Ashish A. Deshmukh, Baojiang Chen, David R. Lairson, Maria Daheri, Sally W. Vernon, Jane R. Montealegre

## Abstract

Home-based self-sample human papillomavirus (HPV) testing may be an alternative for women who do not attend clinic-based cervical cancer screening. We assessed barriers to care and motivators to use at-home HPV self-sampling kits during the COVID-19 pandemic as part of a randomized controlled trial evaluating kit effectiveness. Participants were women, aged 30-65 years and underscreened for cervical cancer in a safety-net healthcare system. We conducted telephone surveys in English/Spanish among a subgroup of trial participants, assessed differences between groups and determined statistical significance at p<0.05. Over half of 233 survey participants reported clinic-based screening (Pap) is uncomfortable (67.8%), embarrassing (52.4%), and discomfort seeing male providers (63.1%). The latter two factors were significantly more prevalent among Spanish versus English speakers (66.4% vs 30% and 69.9 vs 52.2%, respectively, p<0.01). Most women who completed the kit found Pap more embarrassing (69.3%), stressful (55.6%) and less convenient (55.6%) than the kit. The first factor was more prevalent among Spanish versus English speakers (79.6% vs 53.38%, p<0.05). The COVID-19 pandemic influenced most (59.5%) to participate in the trial due to fear of COVID, difficulty making appointments and ease of using kits. HPV self-sampling kits may reduce barriers among underscreened women in a safety-net system.

## Introduction

The disruptions in the US healthcare system due to the COVID-19 pandemic have resulted in a sharp decline in routine primary care, including cervical cancer screening (1). This is expected to lead to gaps in preventive care and increased risk of preventable chronic diseases (2, 3), especially among medically underserved populations. Cervical cancer screening declined by 84% in April 2020 (4), a month after the declaration of the global COVID-19 pandemic, and the rates had not yet fully recovered by June 2021 (5). Prior to the COVID-19 pandemic, racial minorities and those with limited English proficiency were less likely to be screened for cervical cancer compared to their non-Hispanic white and English proficient counterparts (6), leading to disparities in cervical cancer incidence and mortality (7). These populations who were experiencing higher rates of cervical cancer and other chronic illnesses prior to the pandemic are now being faced with widening health disparities due to COVID (8).

Safety net health systems, which provide care regardless of patients’ ability to pay, provide care for a large proportion of the medically underserved population in the US and have become increasingly important during the COVID-19 pandemic (9). The population served by safety net systems predominantly comprises low-income individuals, immigrants, and racial/ethnic minorities (10). These populations are also those disproportionately affected by COVID (11).

Barriers to cervical cancer screening among safety net system patients have not been fully described and thus research to inform targeted approaches to increase screening participation is needed. A previous study found that underscreened women within a safety net system were more likely to have limited knowledge of HPV, and report cost, time and lack of childcare as barriers to Pap screening compared to screened women (12). COVID-19 introduced additional barriers such as fear of contracting the virus and lack of available appointments (13). In this context, alternative screening strategies such as HPV self-sampling may provide opportunities to continue to deliver preventive care. Home-based self-sample human papillomavirus (HPV) testing, whereby women collect their own cervicovaginal sample, has been proposed as a tool to circumvent many of these barriers. Our team is currently evaluating their effectiveness and implementation among underscreened women in a safety net health system. The trial, which began in February 2020, is unique in that data collection has occurred entirely during the COVID-19 pandemic. Here we describe perceived barriers to cervical cancer screening, as well as motivators to use an at-home HPV self-sampling kit during the COVID-19 pandemic among women in an urban safety net health system.

## Materials and Methods

### Participants and Setting

Study participants were part of a larger HPV self-sampling randomized clinical trial, the Prospective Evaluation of Self-Testing to Increase Screening (PRESTIS) study (17). The trial is being conducted in a large, urban safety net health system, Harris Health System, that is 54.1% Hispanic/Latino, 25.9% Black/African American, 11.3% non-Hispanic White, and 8.7% Asian or other (18). The trial began in Feb 2020, paused in March due to COVID-19 related closures, and resumed in August 2020 when COVID-19-related research restrictions were lifted. The trial’s protocol has been described in detail elsewhere (17). Briefly, patients are eligible for PRESTIS if they meet the following inclusion criteria: 1) are 30-65 years of age; 2) have no history of hysterectomy or cervical cancer; 3) are underscreened for cervical cancer (have not had a Pap test in the past 3.5 years or Pap/HPV co-test in the past 5.5 years); 4) have at least 2 visits within the safety net healthcare system in the past 3.5 years; and 5) were currently enrolled in a healthcare coverage or financial assistance plan accepted by the system (including Medicaid/Medicare, private insurance, and county-sponsored coverage). Eligible patients were randomized to one of three study arms: Arm 1) Telephone recall (control) with a reminder to schedule a Pap test; Arm 2) Telephone recall with mailed HPV self-sampling kit (intervention); and Arm 3) Telephone recall with mailed HPV self-sampling kit and an additional reminder/educational call from a health system employee (intervention plus).

### Data Collection

As part of the trial, we conducted a nested survey to assess acceptability and experiences among a subset of randomly selected trial participants randomized to self-sample HPV testing. This study includes telephone survey participants who responded to the survey between August 2020 and September 2022. The survey was administered by trained, bilingual researcher coordinators in the patient’s preferred language (English or Spanish). Participants were asked to provide verbal consent before commencing the survey and were sent a $20 gift card upon completion. This research was reviewed and approved by Baylor College of Medicine and Harris Health System’s Institutional Review Boards (H-44944).

### Measures

The telephone survey was based on a questionnaire used in a previous study (19). Questions assess healthcare access and utilization (including specific questions about experiences during COVID-19-related closures and restrictions), barriers to cervical cancer screening, demographics and telehealth access. Barriers to clinic-based screening were adapted from existing validated instruments (20-22) and assessed using an 18-item scale, with items such as “I don’t have time to get a Pap test” and “It’s difficult to get an appointment for a Pap test.” Responses were on a three-point Likert scale (not at all, a little, very much) with an “unsure/cannot say” option. Motivators were assessed by asking participants who reported using the kit to compare the convenience, stress/anxiety and embarrassment of a Pap and the at-home self-sample kit by selecting whether the Pap at a clinic is more convenient/stressful/embarrassing, the self-sampling kit is more convenient/stressful/ embarrassing or the two screening methods are about the same.

We assessed COVID-related experiences among all survey participants by asking whether the pandemic affected their economic situation, mental health and physical well-being. Responses were on a 3-point Likert scale (large effect, small effect, no effect). To assess the influence of the COVID-19 pandemic, participants who reported using the kit were asked whether the COVID pandemic influenced their decision to participate in the trial. Those who indicated that the pandemic affected their decision were asked the open-ended question “in what way did the COVID-19 pandemic affect your participation in this trial?” The responses were coded using a grounded theory approach after a thorough reading of the recorded responses (23). Codes were then categorized into emerging themes.

### Analysis

Descriptive statistics were used to summarize the data. Chi-square or Fisher’s exact tests for independence were conducted to assess the relationship between survey question responses and demographics. All statistical analyses were conducted using Stata IC 15.

## Results

A total of 233 telephone surveys were completed by patients enrolled in the PRESTIS study between August 2020 and September 2022. Most surveys (61.4%) were conducted in Spanish and most participants (69.5%) were Hispanic/Latino, with the largest proportion (39.4%) born in Mexico (Table 1). Over 95% of participants had a total household income less than $50,000 and 45.6% had less than a high school education.

**Table 1:** Participant characteristics (n=233)

| Patient characteristic |  | N (%) |
| --- | --- | --- |
| Language of Interview | English | 90 (38.6%) |
|  | Spanish | 143 (61.4%) |
| Race/Ethnicity | Hispanic | 162 (69.5%) |
|  | Black/African American | 51 (21.9%) |
|  | White | 14 (6.0%) |
|  | Asian | 3 (1.3%) |
|  | Other | 3 (1.3%) |
| Place of birth | Mexico | 92 (39.7%) |
|  | United States | 81 (34.9%) |
|  | Central America | 48 (20.7%) |
|  | South America | 4 (1.7%) |
|  | Asia | 2 (0.9%) |
|  | Europe | 3 (1.3%) |
|  | Other | 2 (0.9%) |
| Education completed | No formal schooling | 4 (1.7%) |
|  | Some elementary | 15 (6.5%) |
|  | Elementary | 45 (19.6%) |
|  | Some high school | 41 (17.8%) |
|  | High school | 64 (27.8%) |
|  | Some college/vocational school | 33 (14.3%) |
|  | College/vocational school | 28 (12.2%) |
| Household income | <\$10,000 | 27(19.9%) |
| | \$10,000 - \$19,999 | 47 (34.6%) |
| | \$20,000 - \$29,999 | 29 (21.3%) |
| | \$30,000 - \$39,999 | 19 (14.0%) |
| | \$40,000 - \$49,999 | 8 (5.9%) |
| | >\$50,000 | 6 (4.5%) |

### Self-reported barriers

The most commonly reported barriers to cervical cancer screening were a Pap being uncomfortable (67.8%) and being uncomfortable with a male provider (63.1%). More Spanish-speaking participants reported being uncomfortable with a male provider as a barrier (69.9%) (Table 2) compared to English-speaking participants (52.2%, p<0.05). A similar pattern was seen among women who reported that getting a Pap is embarrassing (52.4% overall). Significantly more Spanish-speaking participants reported that getting a Pap test is embarrassing compared to non-Hispanic and English-speaking participants, 66.4% vs. 30%, p<0.01. Most women reported that getting a Pap test was not expensive (68.5%), with significantly more Spanish-versus English-speaking women reporting that getting a Pap is expensive (25.4% vs. 12.2% for English-speaking participants, p<0.05). Most women reported that getting a Pap is uncomfortable (67.8%), and there was no significant difference in the proportions between groups.

**Table 2:**
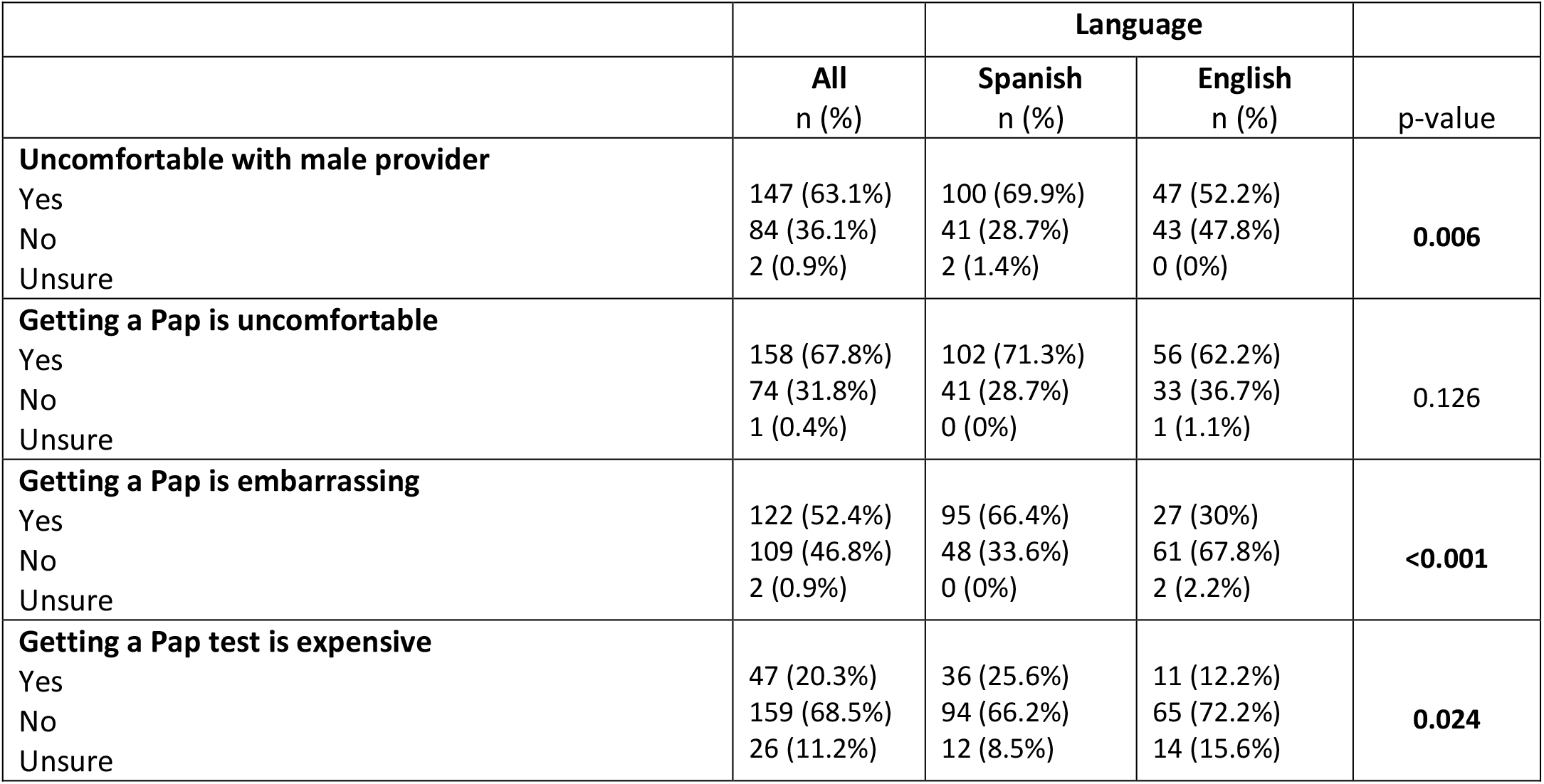
Self-reported barriers (n=233)

|  |  | Language |  |  |
| --- | --- | --- | --- | --- |
|  | All<br>n (%) | Spanish<br>n (%) | English<br>n (%) | p-value |
| **Uncomfortable with male provider** |  |  |  |  |
| Yes | 147 (63.1%) | 100 (69.9%) | 47 (52.2%) | **0.006** |
| No | 84 (36.1%) | 41 (28.7%) | 43 (47.8%) |  |
| Unsure | 2 (0.9%) | 2 (1.4%) | 0 (0%) |  |
| **Getting a Pap is uncomfortable** |  |  |  |  |
| Yes | 158 (67.8%) | 102 (71.3%) | 56 (62.2%) | 0.126 |
| No | 74 (31.8%) | 41 (28.7%) | 33 (36.7%) |  |
| Unsure | 1 (0.4%) | 0 (0%) | 1 (1.1%) |  |
| **Getting a Pap is embarrassing** |  |  |  |  |
| Yes | 122 (52.4%) | 95 (66.4%) | 27 (30%) | **<0.001** |
| No | 109 (46.8%) | 48 (33.6%) | 61 (67.8%) |  |
| Unsure | 2 (0.9%) | 0 (0%) | 2 (2.2%) |  |
| **Getting a Pap test is expensive** |  |  |  |  |
| Yes | 47 (20.3%) | 36 (25.6%) | 11 (12.2%) | **0.024** |
| No | 159 (68.5%) | 94 (66.2%) | 65 (72.2%) |  |
| Unsure | 26 (11.2%) | 12 (8.5%) | 14 (15.6%) |  |

### COVID-related barriers

Most participants who returned the kit (78%) reported that the COVID-19 pandemic affected their economic situation, 46.4% said it affected their mental health, and 39.2% said it affected their physical health. More Spanish-speaking participants reported that COVID-19 related measures affected them economically (82.5%) compared to English-speaking participants (72.2%), though the results were not statistically significant (p=0.052). Conversely, significantly fewer Spanish-speaking participants reported that COVID-19 affected their mental health (37.8%) compared to English-speaking participants (60%, p<0.05). Most participants said that the COVID-19 pandemic did not affect their physical health (60.5%), and there were no significant differences between groups.

### Motivators to participate in self-sample HPV testing

Among participants who reported returning the HPV self-sampling kit, most participants (59.5%) reported that the COVID-19 pandemic influenced their decision to participate in the HPV self-sampling trial (Table 4). The most common reported reasons for why the pandemic influenced the patient’s decision to participate fell into three main categories: fear of getting COVID (41.3%), difficulty getting an appointment (21.7%) and having an easier time completing their screening at home (12%). Other reasons included not having time to travel, having to care for children and having a disability that made going to the clinic difficult. No significant differences in reported reasons were found between language groups.

Most participants who completed self-sampling found the self-sampling kit to be more convenient and less stressful compared to clinic-based cervical cancer screening (both 55.6%), with no significant differences between groups (Table 3). There were no patients who found the self-sampling kit more embarrassing than the Pap test. While most participants found a Pap more embarrassing than the self-sampling kit (69.3%), significantly more Spanish-versus English-speaking participants found the Pap test more embarrassing than using a self-sampling kit (79.6% vs. 53.3%, p<0.05).).

**Table 3:** Motivators-Self-sampling vs. Pap (n=153)

|  |  | Language |  |  |
| --- | --- | --- | --- | --- |
|  | All<br>n (%) | Spanish<br>n (%) | English<br>n (%) | p-value |
| **Convenience of Pap vs. self-sampling** |  |  |  |  |
| Self-sampling more convenient | 85 (55.6%) | 48 (51.6%) | 37 (61.7%) | 0.237 |
| Pap more convenient | 18 (11.8%) | 14 (15.1%) | 19 (31.7%) |  |
| Both are about the same | 50 (32.7%) | 31 (33.3%) | 4 (6.7%) |  |
| **Embarrassment of Pap vs. self-sampling** |  |  |  |  |
| Self-sampling more embarrassing | 0 (0%) | 0 (0%) | 0 (0%) | **0.001** |
| Pap more embarrassing | 106 (69.3%) | 74 (79.6%) | 32 (53.3%) |  |
| Both are about the same | 47 (30.7%) | 19 (20.4%) | 28 (46.7%) |  |
| **Stress/anxiety of Pap vs. self-sampling** |  |  |  |  |
| Self-sampling more stressful | 6 (3.9%) | 4 (4.3%) | 2 (3.3%) | 0.940 |
| Pap more stressful | 85 (55.6%) | 52 (55.9%) | 33 (55%) |  |
| Both are about the same | 62 (40.5%) | 37 (39.8%) | 25 (41.7%) |  |

**Table 4:** COVID-related barriers (n=153)

|  |  | Language |  |  |
| --- | --- | --- | --- | --- |
|  | All<br>n (%) | Spanish<br>n (%) | English<br>n (%) | p-value |
| **COVID-19 affected participation in HPV self-sampling trial** |  |  |  |  |
| Yes | 91 (59.5%) | 58 (62.4%) | 33 (55%) | 0.530 |
| No | 61 (39.9%) | 34 (36.6%) | 27 (45%) |  |
| Don't know | 1 (0.7%) | 1 (1.1%) | 0 (0%) |  |
| **Way that COVID affected participation** |  |  |  |  |
| Fear of getting COVID | 38 (41.3%) | 24 (40.7%) | 14 (42.4%) | 0.304 |
| Difficulties getting appointment | 20 (21.7%) | 16 (27.1%) | 4 (12.1%) |  |
| Easier at home | 11 (12%) | 7 (11.9%) | 4 (12.1%) |  |
| Other | 23 (25%) | 12 (20.3%) | 11 (33.3%) |  |
| **COVID-19 affected economic situation** |  |  |  |  |
| Yes- large amount | 101 (43.4%) | 60 (42%) | 41 (45.6%) | 0.052 |
| Yes- small amount | 82 (35.2%) | 58 (40.6%) | 24 (26.7%) |  |
| No | 50 (21.5%) | 25 (17.5%) | 25 (17.5%) |  |
| **COVID-19 affected mental health** |  |  |  |  |
| Yes- large amount | 34 (14.6%) | 13 (9.1%) | 21 (23.3%) | **0.001** |
| Yes- small amount | 74 (31.8%) | 41 (28.7%) | 33 (36.7%) |  |
| No | 125 (53.7%) | 89 (62.2%) | 36 (40%) |  |
| **COVID-19 affected physical health** |  |  |  |  |
| Yes- large amount | 35 (15%) | 20 (14%) | 15 (16.7%) | 0.633 |
| Yes- small amount | 57 (24.5%) | 33 (23.1%) | 24 (26.7%) |  |
| No | 141 (60.5%) | 90 (62.9%) | 51 (56.7%) |  |

## Discussion

In our assessment of barriers to clinic-based screening during the COVID-19 pandemic, we found that discomfort with the test and with male providers, as well as embarrassment, are important and prevalent barriers to screening among underscreened safety net health system patients. These barriers were more prevalent among women who completed the survey in Spanish. Our results suggest that barriers experienced by women within a safety net healthcare system may differ from those experienced by patients in other health care systems who have difficulty accessing care due to financial reasons and other barriers (6, 14, 15). Similar to other studies conducted in safety net health care systems, we found that additional barriers beyond access and financial barriers, including modesty concerns and discomfort, hinder participation in cervical cancer screening (6, 15).

The motivators for using an at-home HPV self-sampling kit appear to address key barriers to cervical cancer screening found in our survey participants. Most who used the kit found it to be less stressful, embarrassing and more convenient than clinic-based screening. Significantly more Spanish-speaking women found the at-home kits to be less embarrassing than clinic-based screening, a barrier reported significantly more among Spanish-speaking participants. Our findings suggest that self-sampling kits may address or circumvent some the key barriers reported by survey participants within a safety net health system, especially those reported by Spanish-speaking women, and may help to address disparities in cervical cancer screening adherence.

Our results show that COVID-19 was a motivating factor for most respondents to participate in the at-home self-sampling HPV trial and that many patients experienced additional barriers to care since the beginning of the pandemic. The most common reasons for this influence included difficulty making appointments, fear of getting COVID and a broad response that screening was easier at home. While the survey did not probe about this last response, many participants mentioned it in the context of competing priorities amid the pandemic, such as childcare. These responses align with research indicating that the burden of childcare and elder care has fallen disproportionately on women during the pandemic (16).

This study had certain limitations that should be considered when interpreting the results. Because the study was conducted among women in a safety net system that cares for un- and under-insured individuals, results may not be generalizable to women served by other types of health systems. Women in community and other healthcare settings often face significant structural barriers related to access to care due to lack of insurance and/or cost. The prevalent barriers in our study most certainly reflect that financial and insurance barriers are largely removed due to participants’ enrollment in the health system. Additionally, as mentioned, the closed-ended survey format did not allow us to probe on some of the responses, particularly how the COVID-19 pandemic influenced use of the kit. Nonetheless this study is unique in that it gives in-depth insight into the particular barriers experienced by safety net patients during the COVID-19 pandemic. To our knowledge, this is the first analysis of cervical cancer screening barriers among women in a safety net health care system in the COVID-19 era.

In conclusion, mailed at-home HPV self-sampling kits present an opportunity to reduce important barriers to cervical cancer screening among women in a safety net healthcare system. Furthermore, during the COVID-19 pandemic, these barriers were exacerbated by economic, physical, and mental effects of the pandemic. Additional research is needed to understand additional barriers experienced by women during the COVID-19 pandemic and how these might be addressed with new screening tools such as at-home HPV testing using self-sampling.

## Data Availability

The de-identiied data that support the findings of this study are available on request from the corresponding author, Susan Parker. The data are not publicly available due to their containing information that could compromise the privacy of research participants.

## Acknowledgements

The authors would like to thank Harris Health System for their partnership and support throughout the study. This study is supported by a grant from the National Institute for Minority Health and Health Disparities (NIMHD, R01MD013715, PI: JR Montealegre). The NIMHD was not involved in the study design; the collection, analysis, or interpretation of data; the writing of this manuscript; or the decision to submit the manuscript for publication. The REDCap software platform used for data capture is supported by a grant from the National Center for Supporting Translational Sciences (UL1 TR000445).

## Competing Interests

There are no financial or non-financial competing interests for the authors of this manuscript.

## Notes

### Competing Interest Statement

The authors have declared no competing interest.

### Clinical Trial

ClinicalTrials.gov Identifier: NCT03898167

### Author Declarations

This study includes telephone survey participants who responded to the survey between August 2020 and September 2022. The survey was administered by trained, bilingual researcher coordinators in the patient's preferred language (English or Spanish). Participants were asked to provide verbal consent before commencing the survey and were sent a $20 gift card upon completion. This research was reviewed and approved by Baylor College of Medicine and Harris Health System's Institutional Review Boards (H-44944).

